# Mortality from COVID in Colombia and Peru: Analyses of Mortality Data and Statistical Forecasts

**DOI:** 10.1101/2020.08.24.20181016

**Authors:** Patrick E Brown, Zoë R Greenwald, Luis Ernesto Salinas, Gabriel Aguirre Martens, Leslie Newcombe, Hellen Gelband, Jeremy Veillard, Prabhat Jha

**Affiliations:** The Centre for Global Health Research, Unity Health Toronto and Dalla Lana School of Public Health, University of Toronto, Toronto, Ontario, Canada; Department of Statistical Sciences, University of Toronto, Toronto, Ontario, Canada; World Bank, Human Development Network and Colombia Country Office

## Abstract

National predictions of the course of COVID mortality can be used to plan for effective healthcare responses as well as to support COVID policymaking. We developed the Global COVID Assessment of Mortality (GCAM), a statistical model with continually improving precision that combines actual mortality counts with Bayesian inference, to predict COVID trends, currently until December 1, 2020. In Colombia, the GCAM analysis found the peak of COVID mortality around August 12 and an expected total of COVID deaths of 24,000-31,000, or 48%-92% over the total through August 21. In Peru, a first mortality peak occurred around May 24, and given the current trajectory, a second peak is predicted around September 6. Peru can expect 29,000-43,000 COVID deaths, representing an increase of 7%-55% over COVID deaths through August 21. GCAM projections are also used to estimate medical surge capacity needs. To gauge the reliability of COVID mortality forecasts, we compared all-cause mortality from January through June 2020 with average all-cause mortality in previous years in Colombia and Peru, and found that the excesses were consistent with GCAM forecast, most notably a doubling of overall mortality from May 25-June 7^th^ of weeks in Peru. The GCAM results predict that as a percentage of all adult deaths in previous years, Colombia can expect about 13% excess from COVID deaths, whereas Peru can expect 34% excess. Comparisons of GCAM analyses of several other countries with Colombia and Peru demonstrate the extreme variability that characterizes COVID mortality around the world, emphasizing the need for country-specific analyses and ongoing monitoring as more mortality data become available.

## INTRODUCTION

The coronavirus disease (COVID) pandemic has killed over 800,000 people, mostly in high-income countries, and sickened millions more around the world (Basset et al., 2020; Johns Hopkins University & Medicine [JHU], 2020; Worldometers.info, 2020). Mortality, even with acknowledged undercounts (Katz et al., 2020; Montagano, 2020; Office for National Statistics [ONS], 2020; The Economist, 2020), is the least biased metric available to monitor the trajectory of the pandemic, to evaluate the effectiveness of interventions, and to estimate healthcare needs.

Reliable predictions of the COVID burden, using mortality as an indicator, are critical to plan responses to the current and potential future pandemic waves. Here, we determine the excess mortality from COVID in Colombia and Peru by comparing average weekly mortality counts for recent years with 2020 for the weeks corresponding to the first COVID wave. We complement this analysis of recent past trends with forward projections to November 1, 2020 using the latest iteration of the Global COVID Assessment of Mortality (GCAM), an open-source statistical model to project COVID mortality trends. In Colombia and Peru, where the epidemic has not yet crested, we compare the excess in projected all-cause mortality by week and we compare these two countries with results from Italy, where mortality from the first viral wave has peaked; from Florida state in the United States and Spain, both of which show incomplete resolution of the first wave before the start of a second (which we define as a “rebound” mortality peak), and from Australia, where the two separate peaks from the first viral wave are seen arising from two different sub-national viral waves.

For each setting, GCAM uses non-linear regressions to fit an eight-parameter model to the observed daily mortality counts. We used Bayesian inference (Carpenter et al., 2017) to generate several thousand posterior samples of potential epidemic curves for each country. The model builds on observed daily mortality counts for places that have already reached epidemic peaks (such as Hubei, China, which accounted for about 90% of Chinese deaths, and Italy and Spain). For settings where the epidemic is still growing, future death count predictions are necessarily less certain, and GCAM displays a variety of curves that could occur based on Bayesian prior distributions, which are quantifications of expert knowledge of the underlying process and comparisons with other coronaviruses. A semi-automated GCAM website (www.cghr.org/covid) provides regular updates. GCAM is open, transparent, and uses a reasonably simple method that employs publicly-reported mortality data to make plausible projections. The method is designed to improve as mortality data are added daily.

## METHODS

### Excess mortality analysis

We compared excess mortality during the first 26 weeks of 2020 to average mortality rates in previous years for Colombia and Peru. We defined excess mortality as death from all causes minus average expected mortality, following methods reported by Aron et al. (2020).

We calculated the average expected number of deaths in Colombia and Peru from historical data for weekly counts of death. In Colombia, five years of final mortality data were available (2015-2018; Government of Colombia, 2020) and in Peru, three years of final mortality data (2017-2019) were used. The primary method of comparison is graphical analysis of weekly data for weeks 1 through 26. Additionally, we computed the p-score, representing percentage of excess deaths relative to the expected deaths for each week with complete data in 2020. These analyses will be updated as more recent weeks of mortality data are published.

### GCAM Mortality Data Sources

We collected COVID mortality data by age and sex, where reported, from the World Health Organization (WHO), country reports (WHO, 2020a), and from online data collections (Basset et al., 2020; JHU, 2020; Worldometers.info, 2020). We deemed each setting’s epidemic to have begun on the day after the first COVID death was recorded. We forecast deaths until December 1, 2020.

For a comparison with current COVID mortality (mainly from acute pneumonia leading to respiratory failure), we examined the WHO mortality database (WHO, 2018) to estimate average baseline pneumonia and influenza deaths during 2015-17. For the US, we used the Centers for Disease Control and Prevention (CDC) mortality database (National Center for Health Statistics, 2020), and for Russia and China, we used the Global Burden of Disease (Global Burden of Disease Collaborative Network, 2018). We obtained age- and sex-specific population denominators from the United Nations Population Division (United Nations, 2019), and for Hubei, China, and for US states, from national vital statistics (National Bureau of Statistics of China, 2018; National Center for Health Statistics, 2020). We calculated age-specific mortality rates at age 20 years or older for pneumonia and influenza (as COVID deaths are rare in children and adolescents), and present totals for both sexes (Garg et al., 2020).

### Statistical modelling of mortality

GCAM uses Bayesian non-linear regressions to fit a parametric model to the observed daily mortality counts. The form of the model and the prior distributions (which quantify subjective beliefs about the nature of the epidemic) were specified in late March 2020 and informed by the daily COVID mortality counts in Hubei, China and Italy. The daily intensity of death is assumed to follow a skew-normal density function, which is a typical “bell-shaped” normal distribution with an added term to permit an epidemic to rise quickly and fall slowly, as has already been observed with COVID deaths (Basset et al., 2020; JHU, 2020; Worldometers.info, 2020) and with two earlier coronavirus epidemics of Severe Acute Respiratory Syndrome (SARS; WHO, 2003) and Middle East Respiratory Syndrome (MERS; WHO, 2019).

The daily death counts were modelled with a negative binomial distribution, an extension of the more commonly used Poisson distribution but allowing for extra variation via an over-dispersion parameter. The eight parameters estimated for each region cover: total expected deaths, adjusting for age, compared to the peak Italian deaths; peak day; epidemic duration; skewness; over-dispersion; and deaths outside the main epidemic. A few settings, most notably Iran and Florida have death counts that have rebounded following a decline from a first peak, and a second skew-normal curve was added to these epidemic trajectories, bringing the total number of parameters to 14 for these regions.

We used Bayesian inference (Carpenter et al., 2017) to combine prior distributions and observed data, and generated several thousand posterior samples of potential epidemic curves for each country. Bayesian prior distributions are quantifications of expert knowledge of the underlying process. For settings where the epidemic is still accelerating, death counts are less predictive, since a variety of curves can fit any particular series of increasing counts. In these cases, GCAM relies on Bayesian prior distributions. For each country, we present the actual mortality daily totals, projected daily totals, the date of peak mortality, and duration of mortality in excess of baseline years’ pneumonia and influenza totals. We present the most likely pathway and a series of “credible regions” at various probability levels. Credible regions for daily incidence are “central regions” with a specified probability of containing the entire epidemic curve (Myllymäki et al., 2017), whereas the credible regions for cumulative mortality are made up of conventional pointwise intervals.

The Appendix provides full details, including the formula for the model, specifics of the prior distributions for each parameter, and simulated epidemics for a population with Italy’s demographic structure. As validation, we fitted the model to training data of deaths on or before April 22 and compared the projections with data observed up to May 27 for 11 countries or US states (data not shown). The source code is available at www.cghr.org/covid.

## RESULTS

### Excess Mortality Analysis

The excess deaths observed in Colombia and Peru are presented in Figures 1A and 1B, respectively. By sex, deaths were higher among males (Figure 2A and 2B) than females (Figure 2C and 2D) in both countries. In Colombia, the greatest excess mortality occurred during June 15-21 (p-score = 24.8% excess mortality; excess deaths = 1126) and June 22-28 (30.5% excess mortality; excess deaths = 137) (Appendix table 1). These were the most recent data available as of August 15^th^, and indicate that mortality was still rising as of late June. During the initial period of national quarantine (May 24-April 12), all-cause death counts were below that seen in previous years (Appendix table 1 and Appendix Figure 1, March 16-April 12).

**Table 1.** Current COVID deaths, forecasted death ranges, precision estimate, peak death date and ranges, and comparison with current annual pneumonia deaths and days of excess COVID mortality above pneumonia deaths in selected countries.

| Country | Deaths as of Aug. 21 | Forecasted deaths (000s) |  |  | Precision (1 is most) | Initial peak date |  |  | Second peak date |  |  | Max deaths/day |
| --- | --- | --- | --- | --- | --- | --- | --- | --- | --- | --- | --- | --- |
|  |  | Lower | Upper | Median |  | Median | Lower | Upper | Median | Lower | Upper |  |
| Colombia | 16,183 | 24 | 31 | 27 | 1.3 | 30-Apr | 23-Apr | 09-May | 12-Aug | 07-Aug | 19-Aug | 380 |
| Peru | 27,034 | 29 | 42 | 33 | 1.4 | 24-May | 22-May | 26-May | 06-Sep | 18-Aug | 30-Sep | 303 |
| Florida | 10,056 | 12 | 19 | 14 | 1.5 | 23-Apr | 18-Apr | 27-Apr | 11-Aug | 05-Aug | 23-Aug | 284 |
| Spain | 28,813 | 29 | 30 | 29 | 1.0 | 29-Mar | 28-Mar | 30-Mar | 19-Aug | 14-Aug | 30-Aug | 888 |
| Italy | 35,418 | 36 | 36 | 36 | 1.0 | 28-Mar | 27-Mar | 29-Mar | 26-Aug | 21-Aug | 05-Sep | 969 |
| Australia | 472 | 1 | 2 | 1 | 4.4 | 29-Apr | 14-Apr | 13-Jun | 10-Aug | 07-Aug | 07-Sep | 25 |
| **Total** | **117,976** | **131** | **160** | **140** | **1.2** | .. | .. | .. | .. | .. | .. | .. |

Notes: Deaths on August 21 as reported on Coronavirus.app (Basset et al., 2020); precision is the ratio of the lower and upper forecasts. Pneumonia deaths include influenza (International Classification of Diseases or ICD-10 code J10-J11), pneumonia (J12-J18), other acute lower respiratory infections (J20-J22), and, where relevant, SARS (U04).

**Figure 1.**
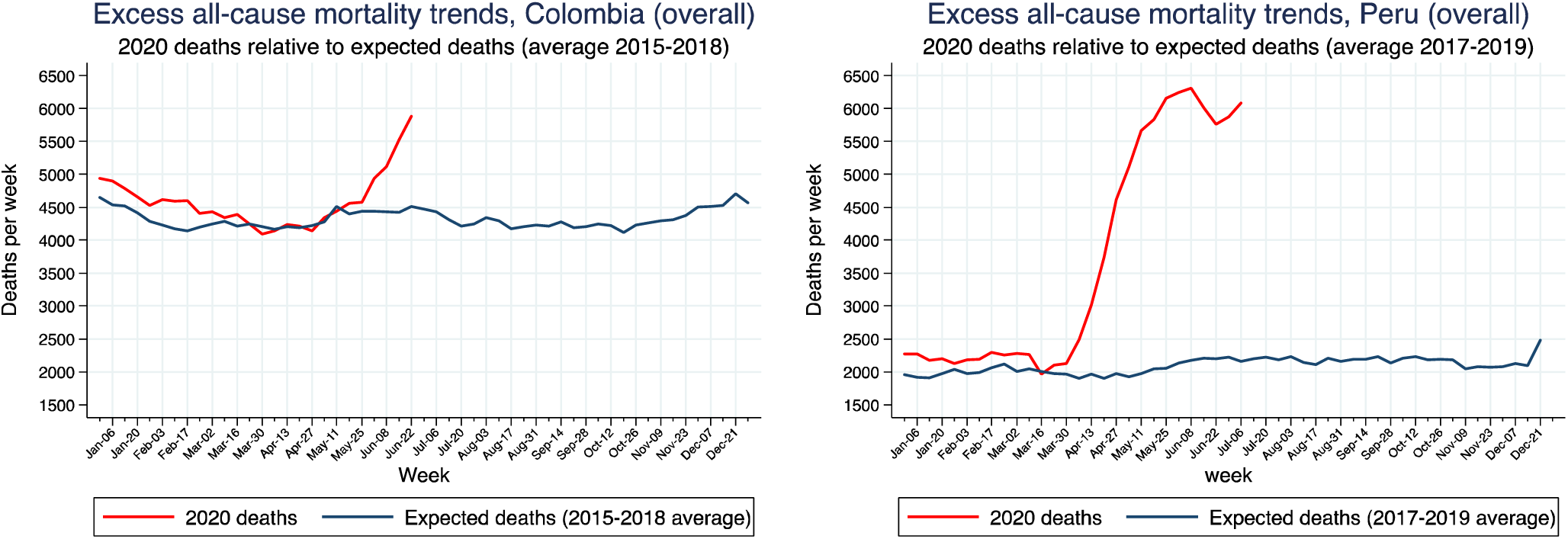
Excess all-cause mortality trends in Colombia (Panel A) and Peru (Panel B)

**Figure 2.**
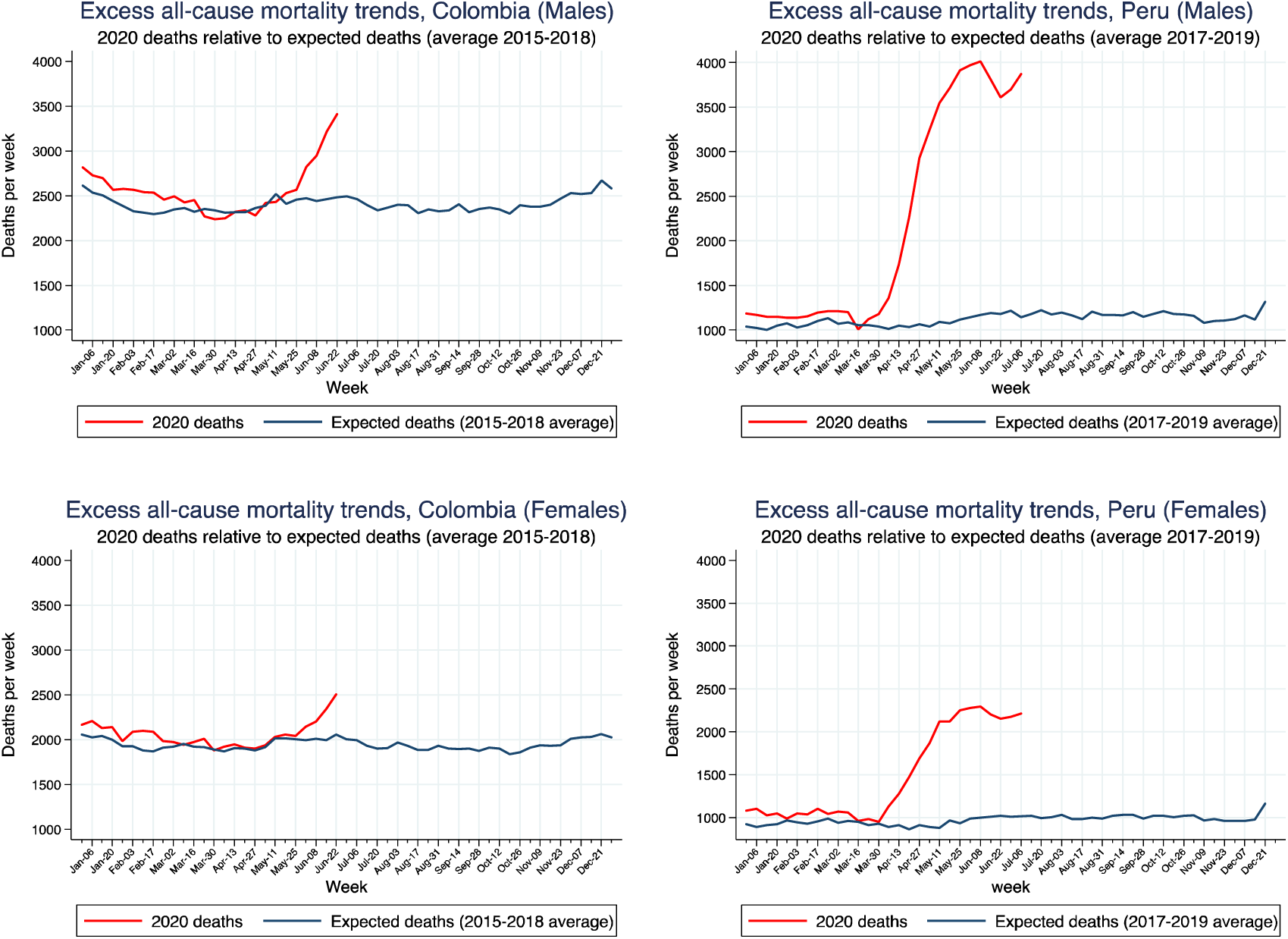
Excess all-cause mortality trends among MALES in Colombia (panel A) and Peru (panel B) and FEMALES in Colombia (panel C) and Peru (panel D)

In Peru, the greatest excess mortality occurred during the weeks of May 25-31 (p-score = 200% excess mortality; excess deaths = 4109) and June 1-7 (p-score = 193% excess mortality; excess deaths = 4115) (Appendix Table 2, Appendix Figure 2).

**Table 2.**
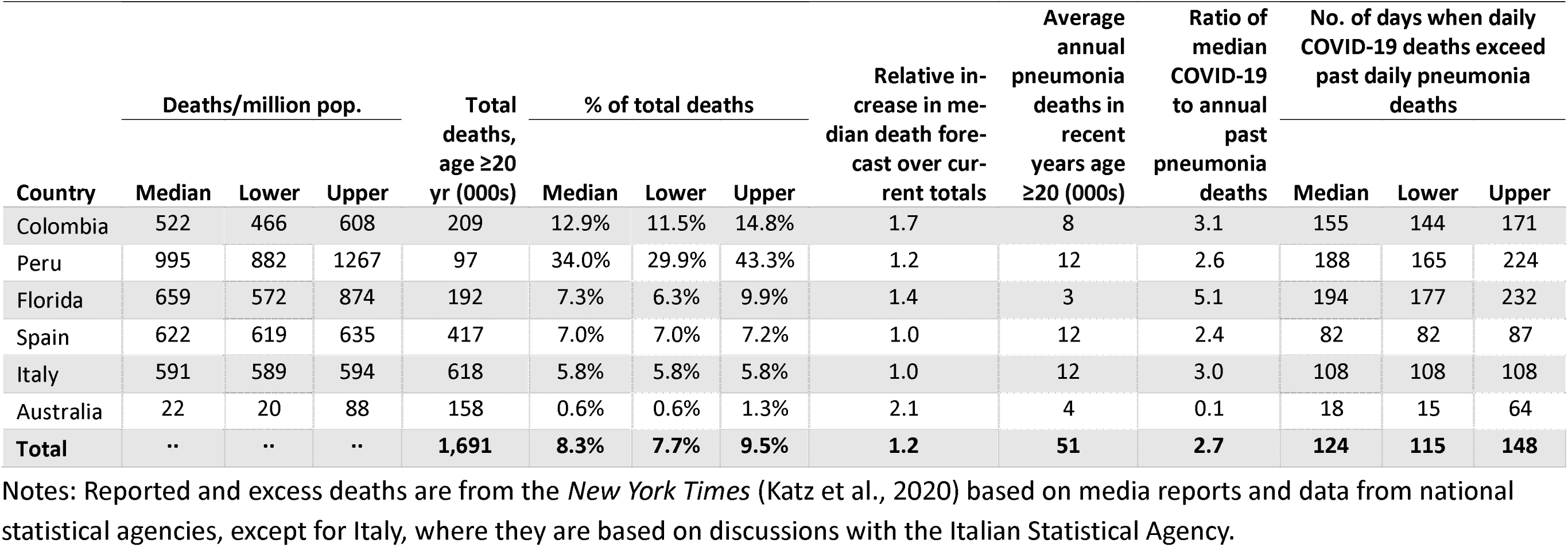
COVID mortality estimates, total mortality, percent of total deaths, and comparison to past pneumonia deaths in selected countries.

### GCAM Analysis

Figure 3 shows the GCAM mortality forecasts for daily COVID deaths until December 1 for Colombia, Peru and for comparison countries with more mature epidemics. The dark line is the median model projection and the red-shaded area represents credible intervals, ranging from 50% (darkest) to 98% (lightest). The actual peak dates are shown for the first or rebound peaks (Australia shows two peaks mostly from different viral peaks in different states). For each graph, the horizontal line represents the uncertainty in the peak date. Colombia’s mortality peaked on August 12 (range August 7-19). Peru’s mortality data show some evidence of a rebound peak, with the first larger peak on May 24 (range May 22-26) and a less certain rebound peak on September 6 (range Aug 18-Sept 30). As comparison, Spain shows a larger first mortality peak on March 29 (range March 28-30) and a smaller, more uncertain peak on August 19 (range August 14-30). Florida shows a larger rebound peak. Italy demonstrates a single peak in late March and Australia shows two distinct peaks. The first represents mostly deaths outside Victoria state and the second peak are deaths within Victoria.

**Figure 3.**
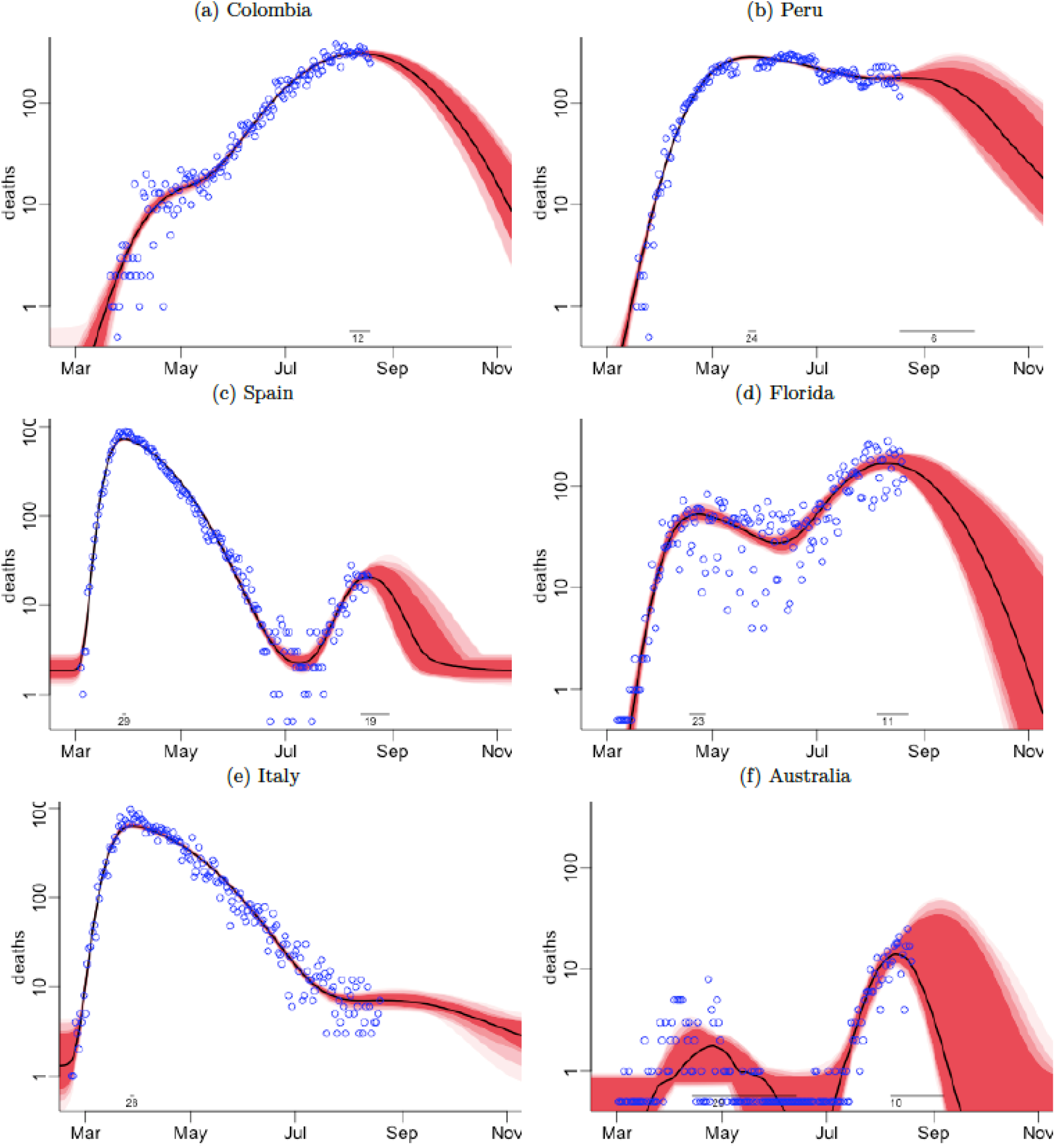
Data and GCAM projections for daily deaths in selected countries. Prediction envelopes (global) are shaded regions, and observed data are blue symbols.

Figure 3 and Table 1 show the range of expected cumulative deaths in the comparison countries. The total number of deaths in Colombia is expected to be 24,000-31,000 (median = 27,000). The lowest value would represent an approximately 48% increase in COVID deaths over totals as of August 21, and the maximum would represent a 92% increase (Table 1). Peru can expect 29,000 to 42,000 deaths (median 33,000) which represents an increase from 7% to 55% above current COVID deaths. We used the average annual deaths in previous years from pneumonia and influenza in both sexes above age 20 years as a crude indicator of historical hospital and ventilator capacity. We compare these totals against the median COVID deaths, and this ratio provides an estimate of the “surge” capacity needed to deal with the excess COVID deaths. This shows that both Colombia and Peru have reasonably high ratios (3.1 and 2.6, respectively) where surge capacity is needed, above the value for Spain (2.4), for example.

For all countries, we estimate an average of 124 days (range: 115-148) until December 1 during which COVID deaths will exceed the daily death total of pneumonia in earlier years (Table 2). For Colombia, this is 155 days (range: 144-171) and for Peru, 188 days (range: 165-224). As a proportion of all annual mortality, the median estimate of excess COVID mortality is 12.9% in Colombia and a very high 34% in Peru.

We compared the GCAM model’s performance to other mortality models. The sixth column in Table 1—”precision”—reports the ratios of upper to lower credible intervals, which is our measure of uncertainty. The precision for Colombia and Peru are reasonably robust at 1.3 and 1.4, respectively. GCAM results showed generally consistent results for the selected countries over the six model runs (Appendix Figure 3). By contrast, results from the Institute for Health Metrics and Evaluation (IHME) were far more uneven, showing lower estimates than GCAM on April 22 and 29, and higher estimates afterwards. Similarly, the IHME median model results indicate that Brazil may have 125,000 deaths by Aug 4^th^ and GCAM estimates an upper limit of 70,000. Moreover, the IHME model does not show consistent improvements in precision as would be expected with more death data for these countries and for others (Appendix Figure 4). Appendix Figure 5 compares GCAM forecasts up to May 27 to IHME forecasts and those produced by the Los Alamos National Laboratory (LANL) COVID-19 Team (2020), training the models on data up to 21 April. Both GCAM and the Los Alamos projections contain the observed mortality data within their credible intervals, but IHME does so less consistently. The credible intervals for the LANL are notably wide.

**Figure 4.**
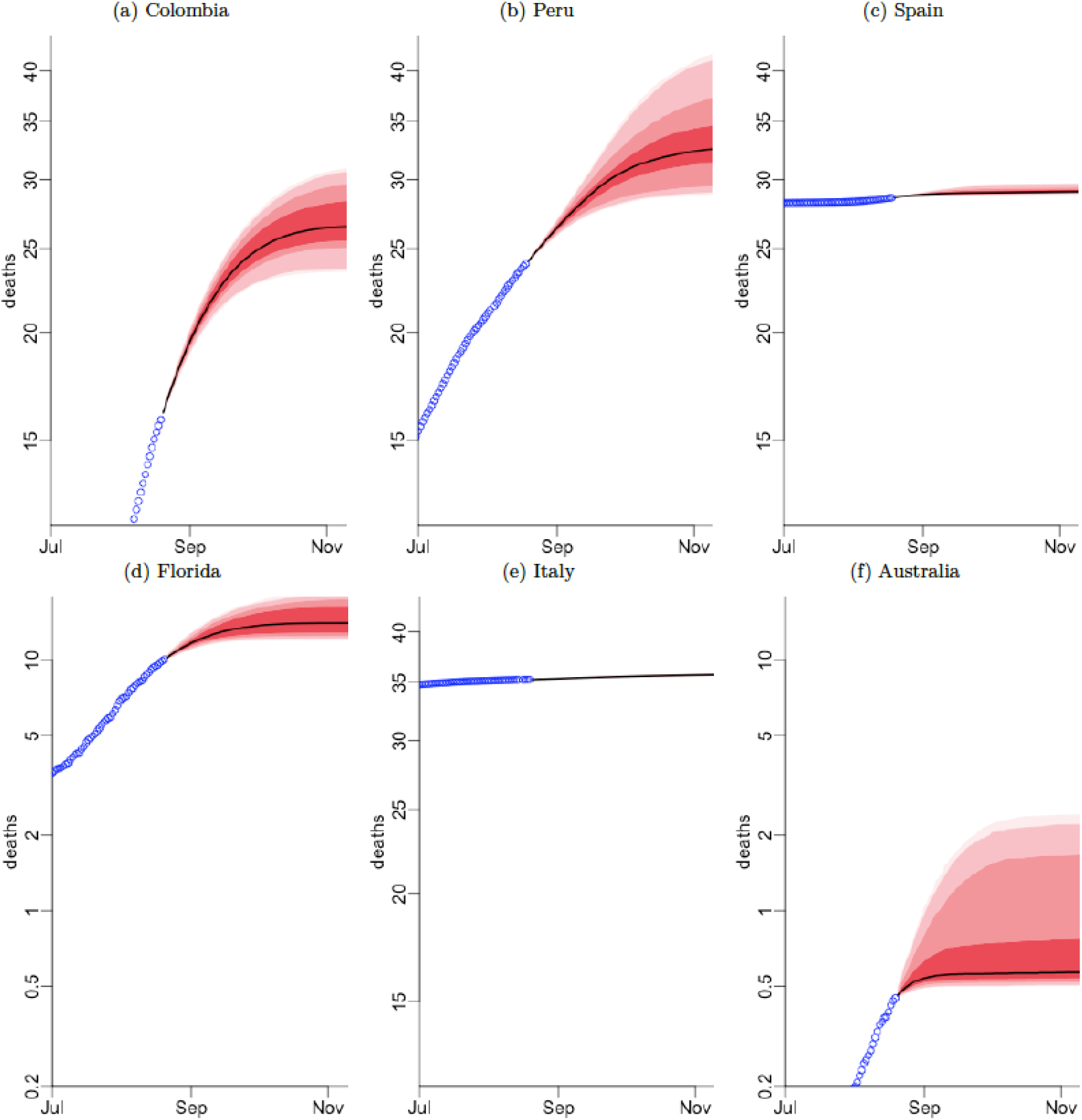
Data and projections for cumulative deaths in selected countries. Prediction envelopes (pointwise) are shaded regions, and observed data are blue symbols.

## DISCUSSION

Colombia and Peru will have a substantial burden of excess deaths due to COVID compared to the deaths in earlier years. Peru’s COVID mortality has already had one peak in late May, and there is the possibility of another peak in early September. Colombia’s COVID mortality peaked in mid-August. In Colombia and Peru, the final total of the first viral wave of COVID deaths will continue to rise by at least 48% in Colombia, and at least 7% in Peru, reaching 24,000 and 29,000 respectively. In the worst case, but still plausible, scenario, totals will reach 31,000 in Colombia and 42,000 in Peru. These totals are broadly consistent with the actual excess mortality from all causes observed in Colombia (30% excess for the week of June 22-28) and in Peru (200% excess for week of May 25-31). Hence, as a proportion of overall adult mortality, the annual excess for the median projection for Colombia is about 13% and for Peru it is 34%. In Colombia, the excess all-cause mortality until July 1, 2020 was most notable among males, and at ages 60 or higher (Government of Colombia, 2020). By contrast, in North America and Europe, most of the excess all-cause and COVID mortality has been at ages 70 or higher. These totals are aggregates, and as recently observed other countries, micro-epidemics with different trajectories are likely to occur, as has been observed in sub-regions of Italy and Spain (Basset et al., 2020; JHU, 2020; Worldometers.info, 2020). Indeed, the two mortality peaks seen in Australia largely represent different epidemics outside and inside the state of Victoria.

An acute excess of hospitalizations and deaths, clustered over short time periods, can swamp any health system. Italy faced more pressures in one month on ventilators and critical-care bed supplies than the usual annual burden (Jha et al., 2020). Estimates for the US suggest that above age 55 years, for every COVID death, ten people will require hospitalization (IHME, 2020). For Colombia and Peru, GCAM provides additional details of when these deaths are likely to peak and the period over which excess deaths will occur, averaging about 163 days in Colombia and 192 days in Peru, but with a wide plausible range. Such information can be used to aid planning and to help avert overwhelming health systems.

Improving the trajectory for COVID mortality in Colombia and Peru depends on the effective and prolonged implementation of non-pharmaceutical interventions such as social distancing and masking. Appendix figures 1 and 2 highlight the COVID excess mortality burden following the implementation of quarantine and suggest that adherence to these measures was suboptimal. Considering the context of COVID transmission and mortality risk within upper middle-income countries such as Peru and Colombia, relative to high-income countries, is also important for understanding the effects of intervention strategies (Walker et al., 2020). For example, while populations tend to be younger in middle-income countries than in high-income countries, elderly populations may face higher COVID attack rates due to larger household sizes with closer intergenerational contact (i.e., elderly individuals in contact with a wider range of age groups), and in particular when nursing homes or long-term care facilities become foci of outbreaks (Walker et al., 2020).

Initial infectious disease models estimated that COVID deaths in the US would reach 1.0-2.2 million (roughly 40%-80% above the usual annual total of 2.8 million deaths) if no preventive action were taken (Ferguson et al., 2020). These same projections suggested 510,000 COVID deaths in the UK, roughly doubling the current annual total of all deaths. To date, the US and UK COVID death totals have been substantially lower, suggesting that these first models were inaccurate (overestimating death rates early on in the epidemic). For similar reasons, these models applied to Colombia and Peru (Walker et al., 2020) are also unreliable. Notably, a key limitation across countries is the absence of population-representative serological surveys on random samples of populations (Mallapaty, 2020) to help establish the true infection-fatality rate. This underscores the need for improved mortality surveillance systems in Colombia and Peru, with the capacity to report mortality data with minimal time lags and by age and place of death (such as long-term care homes). Colombia and Peru would also benefit by launching COVID antibody sero-surveys in the near future to provide nationally representative estimates of the total number of people with a history of infection in each setting. The availability of serological data can substantially enhance the accuracy of modelling, beyond relying on reported cases alone, and can therefore better inform and monitor intervention strategies to minimize harm caused by COVID (Metcalf et al., 2020).

All models, including GCAM, will produce some forecasts that turn out to be wrong. This is due, in part, to undercounts and delayed reporting of COVID deaths, which are now known to be common and variable (Katz et al., 2020; Montagano, 2020; ONS, 2020; The Economist, 2020). For example, the undercount in UK COVID deaths might have been about 40% for some weeks (ONS, 2020). In particular, undercounts in nursing home deaths have emerged as a key issue (Condon et al., 2020). Systematic examination of undercounts across settings is needed to establish if the undercounts have accelerated during the epidemic, which would mean that we have underestimated the speed of the upward trajectory, or if they are more random, in which case our results would be less affected. Future iterations of GCAM will correct for undercounts in all places where they are documented. Definitions of COVID mortality vary across countries. The WHO has created a new International Classification of Diseases, 10th edition (ICD-10) code, U07.2 (WHO, 2020b), but this has been applied inconsistently across countries (Dagblad van het Noorden, 2020). Differences in reporting across countries do not bias the results for any one country, however, as mortality forecasts rely on actual counts within each country. In settings with few deaths over many days, the model becomes highly sensitive to the assumed prior distributions. As with all models, ours emphasizes the need to obtain additional days of observation and rerun the analyses as data improve, and as additional research results inform characteristics of COVID transmission dynamics and the effect of interventions such as physical distancing, focused lockdowns, and the use of masks (Lipsitch et al., 2020).

In a time of crisis, open data sharing is particularly important, hence the full input details and code of GCAM are freely available (www.cghr.org/covid) and we welcome critiques and suggestions on how to refine our estimates. We provide a simple metric of precision that can be used to gauge improvements over time. Future estimates will adopt these methods to examine Asian, African, and Latin American countries, many with weaker health systems and mortality surveillance capacity (Jha, 2014).

## Data Availability

Data used in the GCAM model were taken from publicly available sources (as cited). The GCAM statistical forecasts are updated regularly and made available at http://www.cghr.org/covid.

## Contributors

Conceived the idea and developed the study design: PB, PJ. Data analysis: PB, ZG, LES, LN. Literature review: ZG, LN, PJ. PJ and PB wrote the initial draft, and all authors were involved in commenting on subsequent revisions. PB is the guarantor.

## Acknowledgements

We thank Richard Wen for assistance with the website for data.

## IRB Approval

Not needed as all data are publicly available. No individual patient data were used.

## APPENDIX

**Appendix Table 1:** Excess deaths and excess mortality p-score for Colombia (2020)

| Week start | Week end | Expected deaths<sup>1</sup> | 2020 Deaths | Excess deaths<sup>2</sup> | COVID-19 deaths<sup>3</sup> | Excess mortality P-Score (%)<sup>3</sup> |
| --- | --- | --- | --- | --- | --- | --- |
| 30-Dec-19 | 5-Jan-20 | 4650 | 4942 | 292 | . | 6.3 |
| 6-Jan-20 | 12-Jan-20 | 4536 | 4902 | 366 | . | 8.1 |
| 13-Jan-20 | 19-Jan-20 | 4521 | 4787 | 266 | . | 5.9 |
| 20-Jan-20 | 26-Jan-20 | 4412 | 4660 | 248 | . | 5.6 |
| 27-Jan-20 | 2-Feb-20 | 4285 | 4526 | 241 | . | 5.6 |
| 3-Feb-20 | 9-Feb-20 | 4230 | 4619 | 389 | . | 9.2 |
| 10-Feb-20 | 16-Feb-20 | 4170 | 4591 | 422 | . | 10.1 |
| 17-Feb-20 | 23-Feb-20 | 4141 | 4602 | 461 | . | 11.1 |
| 24-Feb-20 | 1-Mar-20 | 4196 | 4405 | 209 | . | 5.0 |
| 2-Mar-20 | 8-Mar-20 | 4249 | 4435 | 187 | 4 | 4.4 |
| 9-Mar-20 | 15-Mar-20 | 4284 | 4341 | 57 | 2 | 1.3 |
| 16-Mar-20 | 22-Mar-20 | 4216 | 4389 | 174 | 16 | 4.1 |
| 23-Mar-20 | 29-Mar-20 | 4248 | 4249 | 1 | 61 | 0.0 |
| 30-Mar-20 | 5-Apr-20 | 4203 | 4095 | -108 | 124 | -2.6 |
| 6-Apr-20 | 12-Apr-20 | 4162 | 4136 | -26 | 225 | -0.6 |
| 13-Apr-20 | 19-Apr-20 | 4204 | 4238 | 34 | 253 | 0.8 |
| 20-Apr-20 | 26-Apr-20 | 4188 | 4217 | 30 | 278 | 0.7 |
| 27-Apr-20 | 3-May-20 | 4221 | 4142 | -79 | 292 | -1.9 |
| 4-May-20 | 10-May-20 | 4281 | 4340 | 59 | 339 | 1.4 |
| 11-May-20 | 17-May-20 | 4514 | 4440 | -74 | 340 | -1.6 |
| 18-May-20 | 24-May-20 | 4402 | 4560 | 158 | 428 | 3.6 |
| 25-May-20 | 31-May-20 | 4438 | 4574 | 136 | 507 | 3.1 |
| 1-Jun-20 | 7-Jun-20 | 4437 | 4936 | 499 | 682 | 11.2 |
| 8-Jun-20 | 14-Jun-20 | 4432 | 5113 | 682 | 851 | 15.4 |
| 15-Jun-20 | 21-Jun-20 | 4426 | 5523 | 1098 | 1126 | 24.8 |
| 22-Jun-20 | 28-Jun-20 | 4509 | 5882 | 1373 | 1374 | 30.5 |
| **TOTAL** |  | 112,551 | 119,644 | 7094 | 6902 |  |

Notes: (1) Expected weekly deaths estimated using national historic average weekly count of deaths (2015-2018); 2020 death data are preliminary (not yet validated)
(2) Excess deaths are the weekly total of deaths from all causes, minus expected deaths
(3) COVID-19 deaths include confirmed and suspected deaths
(4) P-score represents the percentage of excess deaths relative to expected deaths per week

**Appendix Table 2:** Excess deaths and excess mortality p-score for Peru (2020)

| Week start | Week end | Expected deaths<sup>1</sup> | 2020 Deaths | Excess deaths<sup>2</sup> | COVID-19 deaths<sup>3</sup> | Excess mortality P-Score (%)<sup>3</sup> |
| --- | --- | --- | --- | --- | --- | --- |
| 30-Dec-19 | 5-Jan-20 | 1959 | 2267 | 308 | . | 15.7 |
| 6-Jan-20 | 12-Jan-20 | 1915 | 2273 | 358 | . | 18.7 |
| 13-Jan-20 | 19-Jan-20 | 1911 | 2172 | 261 | . | 13.6 |
| 20-Jan-20 | 26-Jan-20 | 1972 | 2197 | 225 | . | 11.4 |
| 27-Jan-20 | 2-Feb-20 | 2037 | 2124 | 87 | . | 4.3 |
| 3-Feb-20 | 9-Feb-20 | 1975 | 2184 | 209 | . | 10.6 |
| 10-Feb-20 | 16-Feb-20 | 1986 | 2194 | 208 | . | 10.5 |
| 17-Feb-20 | 23-Feb-20 | 2060 | 2296 | 236 | . | 11.5 |
| 24-Feb-20 | 1-Mar-20 | 2120 | 2253 | 133 | . | 6.3 |
| 2-Mar-20 | 8-Mar-20 | 2009 | 2278 | 269 | . | 13.4 |
| 9-Mar-20 | 15-Mar-20 | 2049 | 2260 | 211 | . | 10.3 |
| 16-Mar-20 | 22-Mar-20 | 2003 | 1966 | -37 | 14 | -1.8 |
| 23-Mar-20 | 29-Mar-20 | 1968 | 2100 | 132 | 62 | 6.7 |
| 30-Mar-20 | 5-Apr-20 | 1963 | 2129 | 166 | 175 | 8.5 |
| 6-Apr-20 | 12-Apr-20 | 1902 | 2486 | 584 | 385 | 30.7 |
| 13-Apr-20 | 19-Apr-20 | 1964 | 3012 | 1048 | 669 | 53.3 |
| 20-Apr-20 | 26-Apr-20 | 1900 | 3736 | 1836 | 1053 | 96.7 |
| 27-Apr-20 | 3-May-20 | 1975 | 4618 | 2643 | 1372 | 133.8 |
| 4-May-20 | 10-May-20 | 1928 | 5119 | 3191 | 1543 | 165.6 |
| 11-May-20 | 17-May-20 | 1969 | 5664 | 3695 | 1124 | 187.6 |
| 18-May-20 | 24-May-20 | 2041 | 5831 | 3790 | 768 | 185.7 |
| 25-May-20 | 31-May-20 | 2051 | 6160 | 4109 | 1449 | 200.4 |
| 1-Jun-20 | 7-Jun-20 | 2132 | 6247 | 4115 | 1380 | 193.1 |
| 8-Jun-20 | 14-Jun-20 | 2173 | 6311 | 4138 | 1585 | 190.5 |
| 15-Jun-20 | 21-Jun-20 | 2202 | 6008 | 3806 | 1397 | 172.8 |
| 22-Jun-20 | 28-Jun-20 | 2200 | 5766 | 3566 | 1344 | 162.1 |
| 29-Jun-20 | 5-Jul-20 | 2225 | 5871 | 3646 | 1149 | 163.9 |
| 6-Jul-20 | 12-Jul-20 | 2158 | 6086 | 3928 | 1275 | 182.1 |
| TOTAL |  | 56,745 | 103,608 | 46,863 | 16,744 |  |

Notes: (1) Expected weekly deaths estimated using national historic average weekly count of deaths (2017-2019)
(2) Excess deaths are the weekly total of deaths from all causes, minus expected deaths
(3) COVID-19 deaths include all cases reported by Peru Ministry of Health (MINSa)
(4) P-score represents the percentage of excess deaths relative to expected deaths for each week with complete data in 2020.

**Appendix Figure 1:**
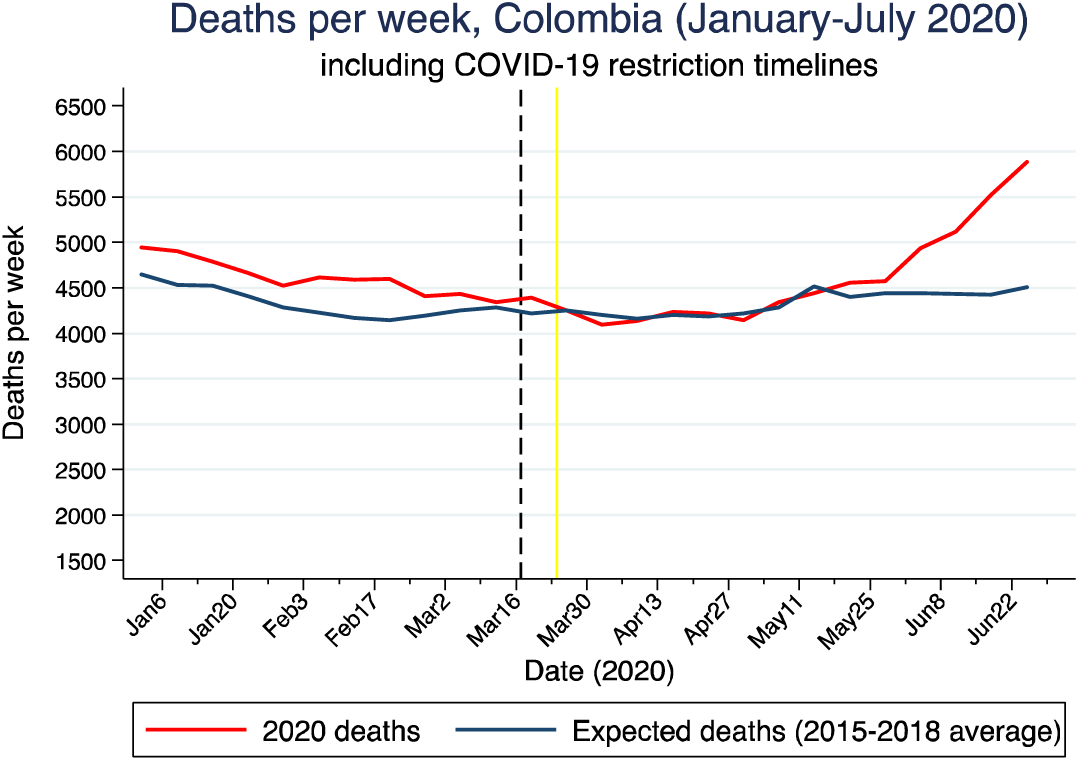
Excess mortality trends in Colombia, 2020 (January – July only), including COVID-19 policies. March 17, 2020 – state of emergency announced (black dashed line) March 24, 2020 – National quarantine begins (end date currently indicated for September 1, 2020)

**Appendix Figure 2:**
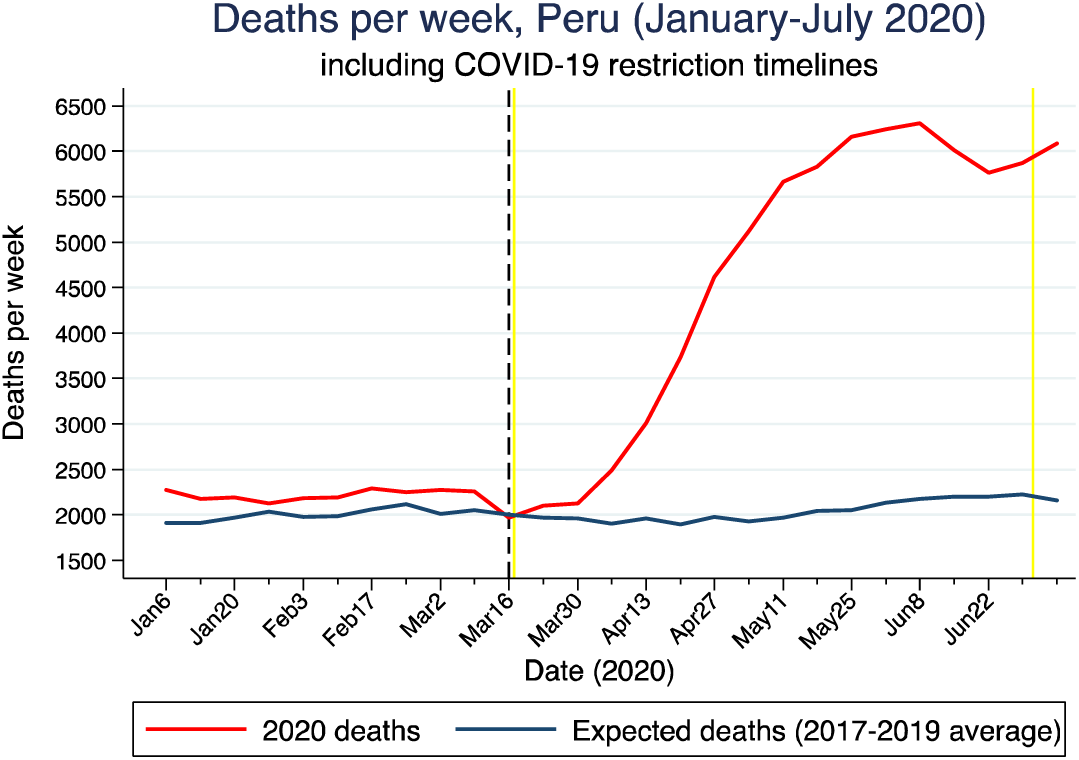
Excess mortality trends in Peru, 2020 (January – July only), including COVID-19 policies. March 15 – State of emergency announced (black dashed line) March 16 – National quarantine begins (first yellow line) July 1 – National quarantine ends (Second yellow line)

### Statistical modelling of mortality

We assumed the mean number of daily deaths for each setting to follow the shape of a skew-normal density function, which resembles a typical “bell-shaped” normal distribution with added skewness to permit an epidemic to rise fast but fall slowly (as has already been observed with COVID-19 deaths, see Basset et al., 2020; Johns Hopkins University, 2020; Worldometers.info, 2020). Each country’s epidemic consists of the sum of two skew normal curves. In some cases (i.e. Australia) the two curves correspond to two separate waves of COVID-19 mortality, whereas in other settings (such as Colombia) the two curves combine to create an epidemic with a single peak. The observed deaths *Y*_*i*_ (*t*) in region *i* on day *t* follows a negative binomial distribution with mean *λ*_*i*_(*t*) and overdispersion parameter *τ*_*i*_, or more specifically:

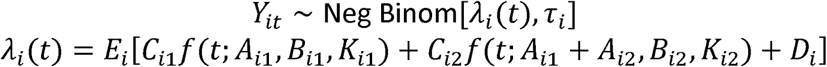

The *E*_*i*_are age and sex standardized death counts, obtained from each country’s population data and age-specific mortality risk calculated from Italian data on 29 March 2020 (EpiCentro, 2020). The model parameters which determine the shape of the epidemic curve are: *C*_*i1*_and *C*_*i2*_, the expected total deaths for each skew-normal curve relative to the Italian rates on 29 March; *A*_*ij*_, the locations of the peaks of the epidemic; *B*_*ij*_, the durations of the epidemic (the skew-normal scale parameters); *K*_*ij*_, the skewness parameters specifying the steepness of the growth phase of the epidemic relative to the decline phase; and *D*_*i*_, a ‘spark’ term allowing for occasional deaths outside the range of the main epidemic.

We used the Markov-Chain Monte Carlo algorithm within the Stan software (Carpenter et al., 2017). From each posterior sample of the model parameters, we computed the intensity *λ*_*i*_ (*t*) and simulated potential future death counts from the negative binomial distribution. We calculated predictions envelopes, which contain the entire true epidemic curve from the samples, with the GET package in R (Myllymäki et al., 2017).

For each country, we present the actual mortality daily totals, projected daily and totals, the date of peak mortality and derive credible intervals (CI), at various probability levels. We calculated age-specific mortality rates at age 20 years or more for influenza and pneumonia, and present totals for both sexes, as COVID-19 deaths are only rarely reported in children and adolescents.

### Prior distributions

We chose Bayesian prior distributions which generate simulated epidemic curves consistent with data from Italy and Hubei, China (Basset et. al, 2020; Johns Hopkins University, 2020; Worldometers.info, 2020), and with SARS and MERS outbreaks (World Health Organization, 2003, 2019). Appendix Table 3 shows the prior distributions for each of the model parameters, the densities for several parameters are displayed in Appendix Figure 3. Figures 3d and 3e show the course of an epidemic for a population of 1 million people with the age distribution of Italy, and the distribution of the final number of total cases appears in 3f. The prior median and 95% prior interval are shown above the horizontal axis of Figure 3f.

The duration parameter *B*_i_ has a 95% prior interval spanning 10 to 35 days, most of a country’s deaths would be expected to occur within a span of four times this parameter. The intensity parameter *C*_*i*_ has a prior which expects the mortality rate in a country with Italy’s age structure to be between 50 and 2,000 per million. The number of deaths per day ranges between 10 and 200 per million at its peak.

**Appendix Table 3:** Prior distributions for model parameters.

| parameter | prior | 95pct interval |
| --- | --- | --- |
| Location $A_{i1}$ | $N(29 \text{ Mar}, 100^2)$ | Apr 2019, Jan 2022 |
| Second location $A_{i2}$ | $\text{Gamma}(3, 0.02)$ | 31, 360 |
| Horizontal scale $B_{ij}$ | $\text{Gamma}(10, 0.5)$ | 9.6, 34 |
| Vertical scale $C_{ij}$ | $\text{Gamma}(1.5, 0.36)$ | 0.3, 13 |
| Skewness $K_i$ | $\text{Gamma}(3, 0.5)$ | 1.2, 14 |
| Spark term $D_i$ | $\text{Exp}(20000)$ | $1.3e-06$, 0.00018 |
| Overdispersion $\tau$ | $\text{Exp}(1)$ | 0.025, 3.7 |

**Appendix Figure 3:**
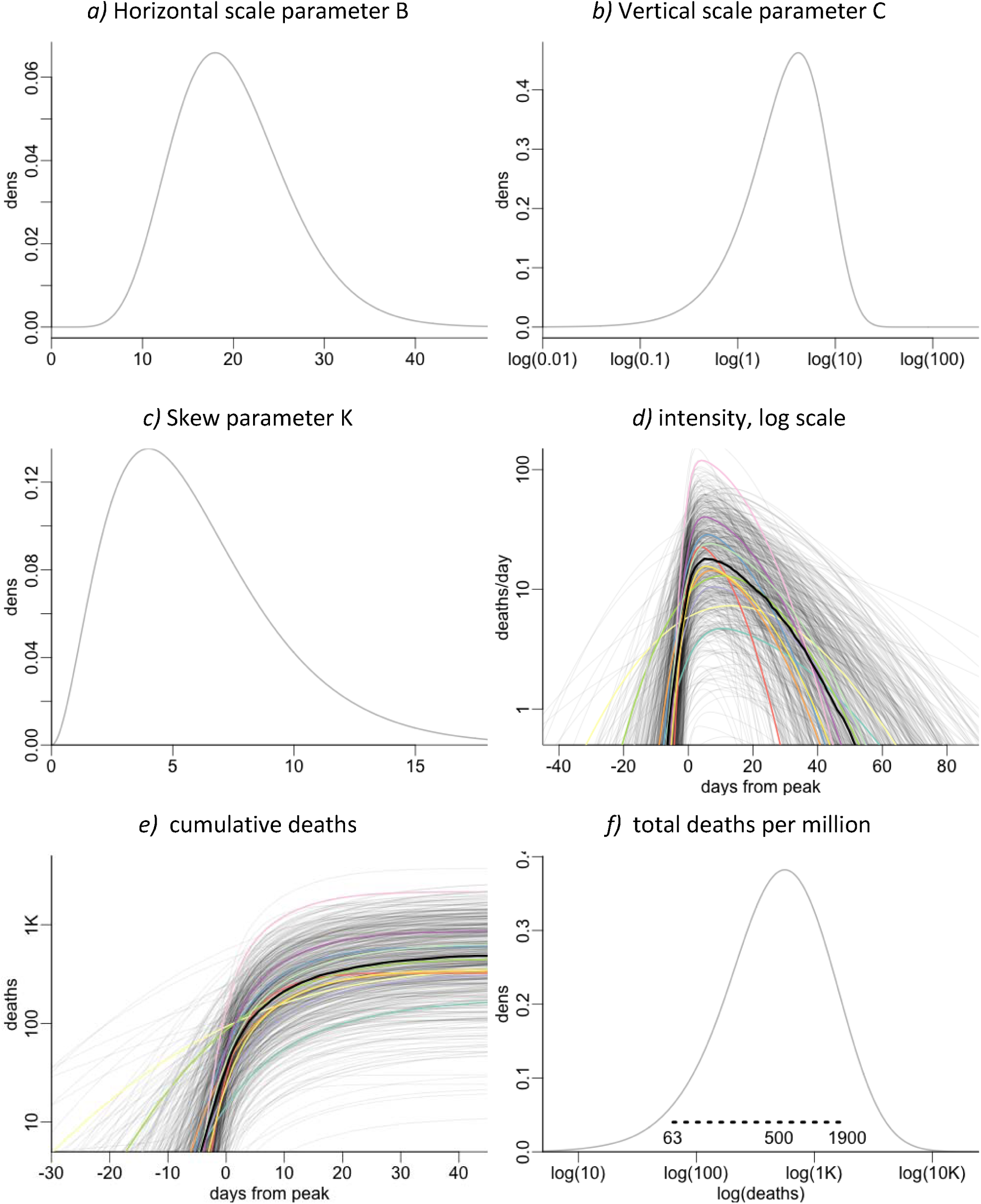
Selected prior distributions, and epidemic trajectories simulated from the priors.

### Validation

As a validation exercise, the model was trained on deaths up to a specified date and predicted the number of deaths expected by 4 August 2020. Six sets of training data were used, with the earliest training period ending on 22 April and the latest training period ending on 26 May. The results were compared to similar forecasts made by IHME (Institute for Health Metrics and Evaluation, 2020) in Appendix Figure 4. As the period of the training data lengthens the uncertainty in the projections decreases, something which is seen most clearly in the predictions for France. Further, while the point estimates change to a certain extent, the prediction intervals are generally overlapping. While the predictions made from the small amount of data available on 22 April are imprecise, the prediction intervals from this date could have been relied upon to encapsulate the eventual predictions made with additional data. The notable exception is Brazil, where predictions on 4 May and earlier are too optimistic.

Appendix Figure 5 compares GCAM forecasts up to May 27 to IHME forecasts and those produced by the Los Alamos National Labs (LANL, 2020), training the models on data up to 21 April. The prediction intervals are 95% pointwise envelopes for IHME and Los Alamos, and an 80% global envelope for the expected number of cases for GCAM. Both GCAM and the Los Alamos projections contain the observed mortality data within their credible intervals, but IHME does so less consistently. The credible intervals for the LANL model are notably wide.

**Appendix Figure 4.**
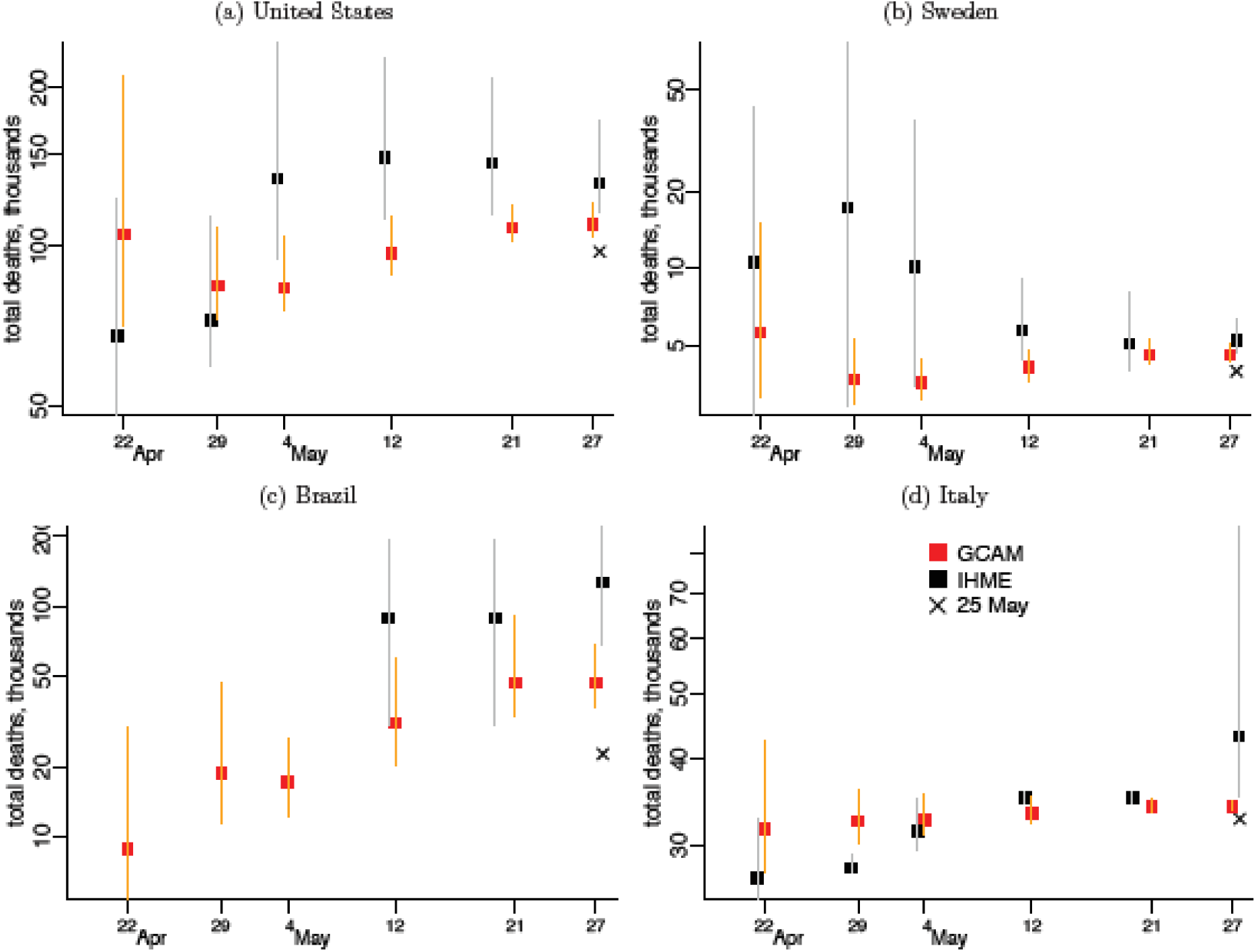
Comparison of six runs of the GCAM model versus the IHME model for selected countries. Forecasted deaths and 95 percent prediction interval for deaths up to August 4 from the GCAM model and from IHME. Six sets of training data were used, with the earliest set of training data consisting of deaths up to April 22 and the latest training data including deaths up to May 26. The “X” represents actual death totals as of May 25.

**Appendix Figure 5:**
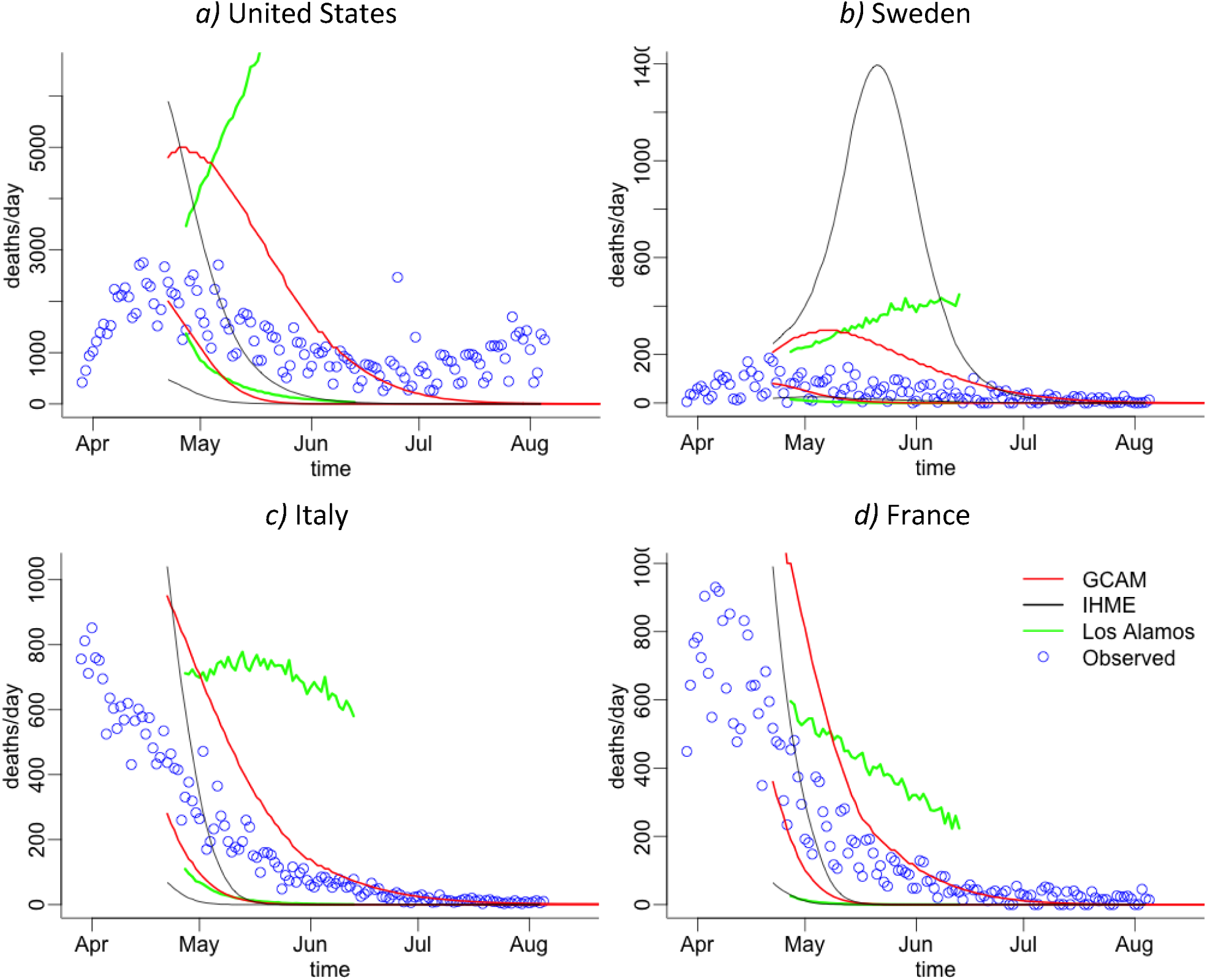
Lower and upper bounds of prediction intervals for daily deaths using training data up to 21 April. Three models are reported: the GCAM model from this paper; the forecasts produced by IHME; and the forecasts from the Los Alamos National Laboratory.

## References

Aron, J., Giattino, C., Muellbauer, J., & Ritchie, H. (2020). A pandemic primer on excess mortality statistics and their comparability across countries. Accessed August 19, 2020, from https://ourworldindata.org/covid-excess-mortality

Basset, K., Michel, M., & Byoun, G. (2020). The coronavirus app. Accessed August 21, 2020, from https://coronavirus.app/map

Carpenter, B., Gelman, A., Hoffman, M. D., Lee, D., Goodrich, B., Betancourt, M., Brubaker, M., Guo, J., Li, P., & Riddell, A. (2017). Stan: A probabilistic programming language. Journal of Statistical Software, 76, 1. https://doi.org/10.18637/jss.v076.i01

Condon, B., Sedensky, M., & Mustian, J. (April 24, 2020). 11,000 deaths: Ravaged nursing homes plead for more testing. AP News. https://apnews.com/e34b42d996968cf9fa0ef85697418b01

Dagblad van het Noorden. (April 3, 2020). CBS: oversterfte van 1200 in laatste week maart. Dagblad van het Noorden. https://www.dvhn.nl/binnenland/CBS-oversterfte-van-1200-in-laatste-week-maart-25528352.html

EpiCentro. (2020). Integrated surveillance of COVID-19 in Italy. Retrieved from https://www.epicentro.iss.it/coronavirus/bollettino/Infografica_29marzo%20ENG.pdf

Ferguson, N., Laydon, D., Nedjati-Gilani, G., Imai, N., Ainslie, K., Baguelin, M., Bhatia, S., Boonyasiri, A., Cucunubá, Z., Cuomo-Dannenburg, G., Dighe, A., Dorigatti, I., Fu, H., Gaythorpe, K., Green, W., Hamlet, A., Hinsley, W., Okell, L. C., van Elsland, S., Thompson, H., Verity, R., Volz, E., Wang, H., Wang, Y., Walker, P. G., Walters, C., Winskill, P., Whittaker, C., Donnelly, C. A., Riley, S., & Ghani, A. C. (2020). Report 9: Impact of non-pharmaceutical interventions (NPIs) to reduce COVID-19 mortality and healthcare demand. Imperial College London. https://www.imperial.ac.uk/media/imperial-college/medicine/mrc-gida/2020-03-16-COVID19-Report-9.pdf

Garg, S., Kim, L., Whitaker, M., O’Halloran, A., Cummings, C., Holstein, R., Prill, M., Chai, S., Kirley, P., Alden, N., Kawasaki, B., Yousey-Hindes, K., Niccolai, L., Anderson, E., Openo, K., Weigel, A., Monroe, M., Ryan, P., Henderson, J., Kim, S., Como-Sabetti, K., Lynfield, R., Sosin, D., Torres, S., Muse, A., Bennett, N., Billing, L., Sutton, M., West, N., Schaffner, W., Talbot, H., Aquino, C., George, A., Budd, A., Brammer, L., Langley, G., Hall, A., & Fry, A. (2020). Hospitalization rates and characteristics of patients hospitalized with laboratory-confirmed coronavirus disease 2019 — COVID-NET, 14 States, March 1–30, 2020. MMWR: Morbidity and Mortality Weekly Report, 69, 458–464. https://doi.org/10.15585/mmwr.mm6915e3

Global Burden of Disease Collaborative Network. (2018). GBD results tool. Institute for Health Metrics and Evaluation. Accessed April 29, 2020, from http://ghdx.healthdata.org/gbd-results-tool

Government of Colombia, Ministry of Health and Social Protection. (2020). Colombia mortality, weeks: 2014-(2Q)2020.

Institute for Health Metrics and Evaluation. (May 26, 2020). COVID-19 projections. Accessed May 27, 2020, from https://covid19.healthdata.org/

Jha, P. (2014). Reliable direct measurement of causes of death in low- and middle-income countries. BMC Medicine, 12(1), 19. https://doi.org/10.1186/1741-7015-12-19

Jha, P., Gelband, H., La Vecchia, C., Qiu, H., Bogoch, I., Brown, P., & Nagelkerke, N. (2020). Reliable quantification of COVID-19 mortality worldwide. OSF Preprints. https://doi.org/10.31219/osf.io/zhwcu

Johns Hopkins University & Medicine. (2020). Mortality analyses - Johns Hopkins Coronavirus Resource Center. Accessed August 21, 2020, from https://coronavirus.jhu.edu/data/mortality

Katz, J., Lu, D., & Sanger-Katz, M. (April 28, 2020). U.S. coronavirus death toll is far higher than reported, C.D.C. data suggests. The New York Times. https://www.nytimes.com/interactive/2020/04/28/us/coronavirus-death-toll-total.html

Lipsitch, M., Swerdlow, D. L., & Finelli, L. (2020). Defining the epidemiology of Covid-19 — Studies needed. New England Journal of Medicine, 382(13), 1194–1196. https://doi.org/10.1056/NEJMp2002125

Los Alamos National Laboratory COVID-19 Team. (2020). COVID-19 confirmed and forecasted case data. Accessed May 28, 2020, from https://covid-19.bsvgateway.org

Mallapaty, S. (2020). Antibody tests suggest that coronavirus infections vastly exceed official counts. Nature, 10.1038/d41586-41020-01095-41580. https://doi.org/10.1038/d41586-020-01095-0

Metcalf, C. J. E., Morris, D. H., & Park, S. W. (2020). Mathematical models to guide pandemic response. Science, 369(6502), 368–369. https://doi.org/10.1126/science.abd1668

Montagano, C. (2020). COVID-19: excess mortality figures in Italy. Towards Data Science. Accessed April 30, 2020, from https://towardsdatascience.com/covid-19-excess-mortality-figures-in-italy-d9640f411691

Myllymäki, M., Mrkvička, T., Grabarnik, P., Seijo, H., & Hahn, U. (2017). Global envelope tests for spatial processes. Journal of the Royal Statistical Society: Series B (Statistical Methodology), 79(2), 381–404. https://doi.org/10.1111/rssb.12172

National Bureau of Statistics of China. (2018). National data: Annual by province. Accessed May 21, 2020, from http://data.stats.gov.cn/english/easyquery.htm?cn=E0103

National Center for Health Statistics. (2020). National Vital Statistics System: Mortality statistics. Accessed May 21, 2020, from https://www.cdc.gov/nchs/nvss/mortality.htm

Office for National Statistics. (2020). Deaths registered weekly in England and Wales, provisional: week ending 10 April 2020. https://www.ons.gov.uk/peoplepopulationandcommunity/birthsdeathsandmarriages/deaths/bulletins/deathsr egisteredweeklyinenglandandwalesprovisional/weekending10april2020

Team, L. A. N. L. C.-1. (2020). COVID-19 Confirmed and Forecasted Case Data. Retrieved from https://covid-19.bsvgateway.org/

The Economist. (April 4, 2020). Fatal flaws: Coronavirus statistics. The Economist, 435(9188), 73. https://www.economist.com/graphic-detail/2020/04/03/covid-19s-death-toll-appears-higher-than-official-figures-suggest

United Nations, Department of Economic and Social Affairs, Population Division. (2019). World Population Prospects 2019. https://population.un.org/wpp/

Walker, P. G. T., Whittaker, C., Watson, O. J., Baguelin, M., Winskill, P., Hamlet, A., Djafaara, B. A., Cucunubá, Z., Olivera Mesa, D., Green, W., Thompson, H., Nayagam, S., Ainslie, K. E. C., Bhatia, S., Bhatt, S., Boonyasiri, A., Boyd, O., Brazeau, N. F., Cattarino, L., Cuomo-Dannenburg, G., Dighe, A., Donnelly, C. A., Dorigatti, I., van Elsland, S. L., FitzJohn, R., Fu, H., Gaythorpe, K. A. M., Geidelberg, L., Grassly, N., Haw, D., Hayes, S., Hinsley, W., Imai, N., Jorgensen, D., Knock, E., Laydon, D., Mishra, S., Nedjati-Gilani, G., Okell, L. C., Unwin, H. J., Verity, R., Vollmer, M., Walters, C. E., Wang, H., Wang, Y., Xi, X., Lalloo, D. G., Ferguson, N. M., & Ghani, A. C. (2020). The impact of COVID-19 and strategies for mitigation and suppression in low- and middle-income countries. Science, 369(i6502), 413–422. https://doi.org/10.1126/science.abc0035

World Health Organization. (2003). Probable cases of SARS by date of onset, worldwide, 1 November 2002 - 10 July 2003. Accessed April 29, 2020, from https://www.who.int/csr/sars/epicurve/epiindex/en/index1.html

World Health Organization. (2018). WHO mortality database. Accessed May 21, 2020, from https://www.who.int/healthinfo/mortality_data/en/

World Health Organization. (2019). Epicurve of confirmed global cases of MERS-CoV. Accessed April 29, 2020, from https://www.who.int/emergencies/mers-cov/epi-16-july-2019.png

World Health Organization. (2020a). Coronavirus disease (COVID-2019) situation reports. Accessed May 21, 2020, from https://www.who.int/emergencies/diseases/novel-coronavirus-2019/situation-reports/

World Health Organization. (2020b). List of official ICD-10 updates. Accessed May 21, 2020, from https://www.who.int/classifications/icd/icd10updates/en/

Worldometers.info. (2020). Worldometer: Coronavirus. Accessed August 21, 2020, from https://www.worldometers.info/coronavirus/

